# Kinetics of SARS-CoV-2 antibody responses pre- and post- COVID-19 convalescent plasma transfusion in patients with severe respiratory failure: an observational case-control study

**DOI:** 10.1101/2020.12.10.20247007

**Authors:** Matthew N. Klein, Elizabeth Wenqian Wang, Paul Zimand, Heather Beauchamp, Caitlin Donis, Matthew D. Ward, Aidaelis Martinez-Hernandez, Ali Tabatabai, John W. Baddley, Evan M. Bloch, Kristin E. Mullins, Magali J. Fontaine

**Affiliations:** Department of Pathology, University of Maryland School of Medicine, Baltimore MD; University of Maryland Saint Joseph Medical Center, Baltimore MD; University of Maryland Medical System, Baltimore MD; University of Maryland Upper Chesapeake Medical Center, Bel Air MD; Department of Medicine, University of Maryland School of Medicine, Baltimore MD; Department of Pathology, School of medicine, Johns Hopkins University, Baltimore, MD

**Keywords:** SARS-CoV-2, antibody, kinetics, convalescent plasma transfusion

## Abstract

**OBJECTIVES:** To investigate if COVID-19 convalescent plasma (CCP) transfusion in patients with severe respiratory failure will increase plasma levels of Severe Acute Respiratory Syndrome Coronavirus 2 (SARS-CoV-2) antibody titers while improving survival and clinical outcomes.

**DESIGN:** Observational, retrospective, control study of anti-Receptor binding domain (RBD) of SARS-CoV-2 IgG and IgM titers from serial plasma samples drawn before and after CCP administration. Clinical improvement in CCP recipients is assessed and compared to COVID-19 control patients.

**SETTING:** Patients hospitalized with severe COVID19, United States, between April 17 and July 19, 2020

**PARTICIPANTS:** 34 patients hospitalized with severe or life threatening COVID-19 and who consented and received a CCP transfusion, 95 control patients with COVID-19 not transfused with CCP. 34 out the 95 control patients were matched for age, sex, and the level of respiratory support required. Patients less than 18 years old were excluded.

**MAIN OUTCOME MEASURES:** Serial trends of anti-RBD of SARS-CoV-2 IgG and IgM titers in CCP recipients are compared to those in control patients. The primary outcome is survival at 30 days, and the secondary outcomes are length of ventilatory and/or extracorporeal membrane oxygenation (ECMO) support, length of stay (LOS) in the hospital, and LOS in the ICU.

**RESULTS:** CCP transfusion occurred in 34 patients at a median of 12 days following COVID-19 symptom onset. Immediately prior to CCP transfusion, patients median anti-RBD SARS-CoV-2 IgG and IgM titers were 1:3200 (IQR, 1:50 to 1:9600) and 1:320 (IQR, 1:40 to 1:640) respectively. Following a Loess regression analysis, the kinetics and distribution of anti-RBD of SARS-CoV-2 IgG and IgM in plasma from CCP recipients were comparable to those from a control group of 68 patients who did not receive CCP. CCP recipients presented with similar survival, similar duration on ventilatory and/or ECMO support, as well as ICU and hospital LOS, compared to a matched control group of 34 patients.

**CONCLUSION:** In the present study, hospitalized COVID-19 patients with severe respiratory failure transfused with CCP presented with high titers of SARS-CoV-2 IgG antibodies before transfusion and did not show improved survival at 30 days.

## INTRODUCTION

The current global health crisis posed by the Severe Acute Respiratory Syndrome Coronavirus 2 (SARS-CoV-2) pandemic demands urgent containment through vaccine development and distribution(1). Pending the arrival of population-based vaccination, the management of Coronavirus Disease 2019 (COVID-19) has nonetheless improved given refined supportive therapies, including hyperoxygenation, steroids, remdesivir, and anticoagulation(2). Another therapy that has been investigated is passive antibody administration through transfusion of convalescent plasma (CCP) (i.e. plasma collected from individuals who have recovered from COVID-19) to prevent the development of severe COVID-19(3). Historically, CCP has been transfused successfully as post-exposure prophylaxis and/or treatment for diverse pathogens, including other coronaviruses (e.g., SARS-1, Middle East Respiratory Syndrome [MERS])(4). Administration of CCP was first attempted during the early stages of the COVID-19 pandemic in China, where it was reported to confer clinical benefit as reflected by faster viral clearance and improved survival(5, 6). Today, over 100,000 patients have been transfused in the United States (US), predominantly through compassionate use programs. The collective findings suggest that CCP is a safe and potentially effective therapy, particularly when administered early and when containing high titer neutralizing antibodies(7, 8).

The sponsoring institution, the University of Maryland Medical Center, has one of the highest acute level care and intensive care unit (ICU) capacities in the US and has been uniquely prepared to treat COVID-19 with different emerging therapies, including CCP. In late March, the United States Food and Drug Administration (FDA) published guidelines for investigational use of CCP, recommending three pathways to access CCP: 1) using an emergency use investigation new drug (IND) application or eIND; 2) using a national expanded access protocol (EAP) centralized by the Mayo Clinic; and /or 3) using a traditional IND to support clinical research trials(3). In partnership with regional referring hospitals, our tertiary care center opted to first use the eIND pathway before transitioning to the EAP. In July 2020, the Mayo Clinic published the EAP preliminary results, citing that the CCP was safe to transfuse and was associated with reduced mortality in patients transfused early after symptom onset compared with patients hospitalized for at least seven days in the ICU(8).

In the current observational study, we evaluated the longitudinal profiles of SARS-CoV-2 antibody titers in plasma from critically ill patients with COVID-19 before and after CCP transfusion and compared them to those measured in control patients not transfused with CCP. Additionally, clinical outcomes of CCP recipients were compared with those from a matched control group.

## METHODS

### Study design

This is an observational retrospective control study to investigate the development of the humoral immune response to SARS-CoV-2 in CCP recipients (n=34) and compare it to the humoral response in a control group of patients not treated with CCP (n=68). A separate comparison of clinical outcomes is performed between CCP recipients and a matched control group of patients untreated (n=34). This study was approved by the University of Maryland Baltimore Institutional Review Board (HP-00092606).

### CCP treated subjects

Patients considered for enrollment in the study presented with severe COVID-19 and were hospitalized at three different University of Maryland Medical System Hospitals, University of Maryland University Medical Center (n=23), University of Maryland St. Joseph Medical Center (n=4), University of Maryland Upper Chesapeake Medical center (n=7). Patients were then evaluated for CCP transfusion by an infectious disease clinician based on FDA recommended guidance (https://www.fda.gov/regulatory-information/search-fda-guidance-documents/investigational-covid-19-convalescent-plasma)(3). An institutional ethics committee reviewed the indication of each CCP transfusion. Patients less than 18 years old were excluded. Informed consent was obtained, and CCP was transfused following FDA authorization through either the eIND pathway or the Mayo Clinic EAP. All CCP transfusions occurred between April 17 and July 19, 2020, in patients with a confirmed laboratory diagnosis of COVID-19 and presenting with severe or life threatening COVID-19(7). CCP units with a SARS-CoV-2 antibody titer>1:160, per FDA guidance, were procured by the regional blood center following donor collection qualification (3). Following transfusion, CCP recipients were closely monitored for a minimum of four hours for possible transfusion-related adverse events (TRAE). Blood samples for SARS-CoV-2 antibody titers were collected at specific time points: immediately pre-transfusion (day 0) and days 3, 7, and 14 post-transfusion. Data from three of the CCP recipients were excluded from the kinetics analysis due to insufficient plasma sample quantity; these were still included in the clinical outcome analysis.

### Control subjects

A separate control group (**Control A**, n=68) of hospitalized COVID-19 patients at University of Maryland University Medical Center, who did not receive CCP, were evaluated. Remnant plasma samples from these control patients were aliquoted 1-3 days following collection and stored at −70°C prior to antibody measurement. Sample draws from these patients ranged from 0-48 days after the onset of symptoms. Symptoms in these patients varied from asymptomatic to patients requiring extracorporeal membrane oxygenation (ECMO) support.

To investigate the independent effect of CCP therapy on patients’ clinical outcomes with COVID-19, CCP recipients were retrospectively matched to COVID-19 patients admitted in the same hospital but who did not receive CCP as part of their therapeutics. These control patients (**Control B**, n=34) were matched based on sex, age, and on three levels of respiratory support requirement (non-ventilated, mechanically ventilated, and ventilated with ECMO). Patients who were administered CCP at an outside institution prior to their admission, pregnant, or had instructions not to escalate care (DNI/DNR) were excluded. Seven controls were included in both controls A and control B.

### Clinical data collection and outcomes

After enrollment, the following clinical variables were collected from electronic medical records for CCP recipients and control patients: symptoms at presentation, the level of respiratory support (mechanical ventilation/ECMO status), comorbidities, inflammatory marker plasma concentrations (C-reactive protein (CRP), ferritin, d-dimer, and fibrinogen), other SARS-CoV-2 directed therapies, 30-day in-hospital mortality, number of days on mechanical ventilation, number of days on ECMO support, ICU length of stay (LOS), and hospital LOS.

Clinical improvement was assessed primarily on survival at 30 days. Secondary outcomes included the number of days on ventilatory and/or ECMO respiratory support, LOS in the hospital, and the ICU.

### SARS-CoV-2 receptor binding domain production

The SARS-CoV-2 spike protein receptor-binding domain (RBD) used for the IgM and IgG enzyme-linked immunosorbent assays (ELISAs) were produced following the protocol outlined by Stadlbauer et al., 2020(9). In brief, plasmids containing the SARS-CoV-2 spike protein RBD (provided by the Krammer Laboratory, Icahn School of Medicine at Mount Sinai, NY) were transformed into competent *E. coli* (New England BioLabs; Ipswich, MA) and grown in Luria-Bertani broth with ampicillin overnight in shaker flasks. Plasmids were purified using the Purelink Hipure Plasmid Filter Maxiprep Kit (Invitrogen; Carlsbad, CA). Purified plasmids were transfected, and protein was produced in Expi293 Cells using the Expi293 expression system (Gibco Laboratories; Gaithersburg, MD) per manufacturer instructions. Protein was purified using Ni-NTA resin (Qiagen; Hilden, Germany) and 30 kDa Amicon Ultra Centrifugal Filter Units (MilliporeSigma; Burlington, MA). Protein concentration was measured by NanoDrop (Thermo Fisher Scientific; Waltham, MA), and SDS-PAGE assessed purity.

### Anti-RBD ELISA

Detection of anti-RBD IgG and IgM was completed using an in-house ELISA based on the previously described assay (9). ELISA plates (Thermo Fisher Scientific) were pre-coated overnight with RBD. Plates were washed, blocked, and washed again before an eight-step, four-fold serial dilution (starting at 1:100 for IgG or 1:40 for IgM) of plasma samples were added and incubated for one hour. The wells were then washed, incubated for one hour with either Horseradish Peroxidase conjugated goat-anti-human IgG or IgM detection antibody (1:12000) (Invitrogen), washed, incubated with 3,3′,5,5′-Tetramethylbenzidine (TMB) substrate (Seracare; Milford, MA) for 10 minutes in the dark, and quenched with 1N sulfuric acid (Thermo Fisher Scientific). Plates were then immediately read at an absorbance of 450 nm. Seroconversion was defined as any measurement of anti-RBD IgG or IgM of greater than or equal to 1:100 (IgG) or 1:40 (IgM) titers.

Samples collected from patients prior to the COVID-19 pandemic (collected in 2012) served as negative controls, while plasma samples from individuals with PCR confirmed SARS-CoV-2 infections served as positive controls. Negative controls along with three dilutions of pooled plasma from positive controls were measured on all plates to ensure consistency across all runs. The specificity was evaluated using 45 and 32 plasma samples from negative controls for the anti-RBD IgG and anti-RBD IgM ELISAs, respectively. Twenty-four positive control samples were also measured on the Ortho VITROS total anti-SARS-CoV-2 Ig platform.

### Statistical analysis

Plots of trends on antibody titers vs. the number of days post symptom onset were evaluated using Loess regression analysis with a span of 0.75 and 95% confidence intervals. Statistical significance for categorical variables (e.g., sex, comorbidities, symptoms at presentation, disease severity, ABO, and other SARS-CoV-2 directed therapies) was determined using Fisher’s exact test due to relatively small sample sizes in each group. Quantile-quantile and density plots were examined, and Shapiro-Wilk tests were conducted for the continuous covariates in Table 1 to determine if the assumption of normality was valid. Welch’s one-way ANOVA was used to determine the statistical significance of the normally distributed continuous age variable, and the Kruskal-Wallis was used to determine the statistical significance of the non-normal baseline inflammatory marker concentrations. Odds ratios with Wald confidence intervals and p-values were used to compare CCP recipients and matched controls on changes in inflammatory marker concentrations, the number of days mechanically ventilated or on ECMO support, ICU LOS, and hospital LOS. To compare the 30-day in-hospital mortality of CCP recipients and controls, Kaplan Meier curves were used. The curves ceased at 42 days POS because the median day of transfusion including the CCP recipients who had insufficient plasma samples for the kinetics analysis was 12.5 days POS. A log-rank test was used to determine statistical significance between the survival distribution of CCP recipients and matched controls and is depicted on the Kaplan-Meier curves. An alpha value of 0.05 or less was considered statistically significant. Statistical analysis was performed using R statistical software (Foundation for Statistical Computing; Vienna, Austria) and Prism 8 (GraphPad; San Diego, CA).

**Table 1.** Demographics and clinical characteristics.

|  | **Overall**<br>(n=129) | **CCP Group**<br>(n=34) | **Control A**<br>(n=68) | **Control B**<br>(n=34) | **p-value**<br>CCP vs A | **p-value**<br>CCP vs B |
| --- | --- | --- | --- | --- | --- | --- |
| Male sex, n (%)<sup>1</sup> | 88 (68.2) | 23 (67.6) | 46 (67.6) | 23 (67.6) | 1 | 1 |
| Age (years), mean (SD) <sup>2</sup> | 57.4 (16.4) | 55.4 (16.6) | 59.0 (16.8) | 57.2 (15.3) | 0.31 | 0.65 |
| **Comorbidities, n (%)<sup>1</sup>** |  |  |  |  |  |  |
| BMI >30 | 25 (19.4) | 6 (17.6) | 18 (26.5) | 4 (11.8) | 0.46 | 0.73 |
| Diabetes | 50 (38.8) | 14 (41.2) | 28 (41.2) | 12 (35.3) | 1.00 | 0.80 |
| Hypertension | 67 (51.9) | 16 (47.1) | 38 (55.9) | 17 (50.0) | 0.41 | 1.00 |
| Chronic Obstructive<br>Pulmonary Disease | 10 (7.8) | 3 (8.8) | 7 (10.3) | 2 (5.9) | 1.00 | 1.00 |
| Chronic Kidney Disease | 14 (10.9) | 2 (5.9) | 11 (16.2) | 1 (2.9) | 0.21 | 1.00 |
| Hyperlipidemia | 36 (27.9) | 7 (20.6) | 27 (39.7) | 4 (11.8) | 0.07 | 0.51 |
| Coronary Artery Disease | 8 (6.2) | 1 (2.9) | 6 (8.8) | 3 (8.8) | 0.42 | 0.61 |
| Other Therapies | 2 (1.6) | 2 (5.9) | 0 (0.0) | 0 (0.0) | 0.11 | 0.49 |
| **Symptoms, n (%)<sup>1</sup>** |  |  |  |  |  |  |
| Dyspnea | 118 (91.5) | 32 (94.1) | 59 (86.8) | 33 (97.1) | 0.33 | 1.00 |
| SpO<sub>2</sub><93% | 107 (82.9) | 28 (82.4) | 51 (75.0) | 34 (100.0) | 0.46 | 0.03 |
| Respiratory Rate >30 | 74 (57.4) | 19 (55.9) | 33 (48.5) | 24 (70.6) | 0.53 | 0.31 |
| Arterial O<sub>2</sub>/FiO<sub>2</sub><300 | 107 (82.9) | 34 (100.0) | 45 (66.2) | 33 (97.1) | <0.001 | 1.00 |
| Respiratory Failure | 109 (84.5) | 34 (100.0) | 47 (69.1) | 34 (100.0) | <0.001 | 1.00 |
| Septic Shock | 48 (37.2) | 15 (44.1) | 25 (36.8) | 13 (38.2) | 0.52 | 0.81 |
| **Disease Severity n (%)<sup>1</sup>** |  |  |  |  | 0.001 | 1 |
| Non-ventilated | 39 (30.2) | 6 (17.6) | 29 (42.6) | 6 (17.6) |  |  |
| Mechanical Ventilation | 65 (50.4) | 17 (50.0) | 35 (51.5) | 17 (50.0) |  |  |
| ECMO | 25 (19.4) | 11 (32.4) | 4 (5.9) | 11 (32.4) |  |  |
| **Inflammatory Markers<sup>3</sup>**<br>**median concentration, [IQR]** |  |  |  |  |  |  |
| Day 0 CRP (mg/dL) | 17.5 [6.6, 28.3] | 17.6 [5.4, 32.0] | NA [NA, NA] | 17.5 [13.1, 20.1] | NA | 0.85 |
| Day 0 D-dimer (ng/mL) | 3305.0 [1708.0,<br>7332.5] | 3235.0 [1588.0,<br>6691.3] | NA [NA, NA] | 5215.0 [2192.5,<br>9672.5] | NA | 0.43 |
| Day 0 Ferritin (ng/mL) | 700.6 [445.7,<br>1,116.4] | 924.8 [534.6, 1952.6] | NA [NA, NA] | 494.3 [427.5,<br>731.5] | NA | 0.16 |
| Day 0 Fibrinogen (mg/dL) | 598.00 [484.0, 776.8] | 653.0 [541.0, 800.0] | NA [NA, NA] | 546.0 [385.0,<br>653.0] | NA | 0.09 |
| **ABO, n (%)<sup>1</sup>** |  |  |  |  | <0.001 | 0.002 |
| A neg | 4 (3.1) | 0 (0.0) | 3 (4.4) | 2 (5.9) |  |  |
| A pos | 30 (23.3) | 6 (17.6) | 17 (25.0) | 9 (26.5) |  |  |
| AB pos | 2 (1.6) | 0 (0.0) | 2 (2.9) | 0 (0.0) |  |  |
| B neg | 1 (0.8) | 0 (0.0) | 1 (1.5) | 0 (0.0) |  |  |
| B pos | 20 (15.5) | 8 (23.5) | 10 (14.7) | 2 (5.9) |  |  |
| O neg | 1 (0.8) | 0 (0.0) | 1 (1.5) | 0 (0.0) |  |  |
| O pos | 45 (34.9) | 20 (58.8) | 14 (20.6) | 13 (38.2) |  |  |
| N/A | 26 (20.2) | 0 (0.0) | 20 (29.4) | 8 (23.5) |  |  |
| **Other therapies, n (%)<sup>1</sup>** |  |  |  |  |  |  |
| Hydroxychloroquine | 70 (54.3) | 18 (52.9) | 39 (57.4) | 19 (55.9) | 0.68 | 1.00 |
| Azithromycin | 54 (41.9) | 9 (26.5) | 38 (55.9) | 12 (35.3) | 0.01 | 0.60 |
| Steroids | 21 (16.3) | 11 (32.4) | 2 (2.9) | 8 (23.5) | <0.001 | 0.59 |
| Tocilizumab | 30 (23.3) | 14 (41.2) | 10 (14.7) | 7 (20.6) | 0.01 | 0.11 |
| Remdesivir | 25 (19.4) | 7 (20.6) | 11 (16.2) | 7 (20.6) | 0.59 | 1 |
| Stem Cells | 4 (3.1) | 0 (0.0) | 3 (4.4) | 2 (5.9) | 0.55 | 0.49 |

SD: Standard deviation; IQR: Inter-quartile range; CCP: Convalescent Plasma; ECMO: Extracorporeal membrane oxygenation.
Analysis of statistically significant differences between groups performed using <sup>1</sup>Fisher's Exact Test, <sup>2</sup>Welch's One-Way ANOVA,<sup>3</sup>Kruskal-Wallis Test.

### Patient and public involvement

Neither patients nor the public were involved in our research’s design, conduct, reporting, or dissemination plans.

## RESULTS

### Validation of Anti-RBD IgG and IgM ELISAs

Negative control samples screened by the anti-RBD IgG and IgM ELISAs were found to have a mean optical density (OD) of 0.184 ± 0.127 and 0.222 ± 0.107, respectively. Samples were considered positive for SARS-CoV-2 antibodies if the screening dilution OD was higher than 0.566 for the anti-RBD IgG ELISA and 0.544 for the anti-RBD IgM ELISA.

Of the 24 positive controls screened on both ELISAs as well as the Ortho VITROS platform, 22 patients were positive for anti-RBD IgG antibodies, and 23 patients were positive for anti-RBD IgM antibodies. The median anti-RBD IgG antibody titer was 1:6400, and the median anti-RBD IgM antibody titer was 1:240. Twenty-two (22) of the 24 patients had detectable SARS-CoV-2 antibodies when measured with the Ortho VITROS platform, with a median signal to a cut-off ratio (S/C) of 490. Two patients with IgM titers of 1:40 and undetectable IgG titers were negative for total SARS-CoV-2 antibodies by the Ortho VITROS method. (**Supplemental Figure 1**)

### Characteristics of CCP recipients compared to controls

CCP transfusion was considered and reviewed by a clinician, infectious disease expert, for 41 COVID-19 patients, of whom 34 patients were transfused with CCP upon obtaining consent. Reasons for non-transfusion included patients or legally authorized proxy changing their mind about the treatment. The anti-RBD IgG and anti-RBD IgM responses of these CCP recipients were compared to those of 68 non-transfused control patients (Control A); CCP recipients presented with more severe disease requiring ECMO support, but both groups were similar in terms of sex and age (**Table 1**).

CCP recipients and matched controls (Control B) had similar frequencies of comorbidities, symptoms, and inflammatory marker concentrations at presentation (**Table 1**). Other COVID-19 directed therapies administered during hospitalization were similar as well. ABO type distribution was different between the groups, although it was not available on eight of the 34 (23.5%) matched control patients (**Table 1)**.

### Kinetics of anti-RBD IgG and IgM responses are similar in CCP recipients and control patients

Anti-RBD IgG and IgM responses were examined based on titer levels measured in plasma samples drawn on successive days post-onset of symptoms (POS), starting on the day of transfusion in CCP recipients, which was on a median day 11 POS (IQR, 7.5 to 16.5), and a median day 9.5 POS (IQR, 5 to 17.3) in control patients.

The frequency of patients□who generated an anti-RBD IgG and/or an IgM response was similar□in CCP recipients compared to controls□ (Frequency of IgG response: 100%□(31/31)□and□100%□(68/68) (Frequency of IgM response: 96.8%□(30/31)□and□100%□(68/68)). Furthermore, the seroconversion rate for both anti-RBD IgG and IgM responses, analyzed using a cumulative frequency plot, was similar in CCP recipients compared to controls (**Figures 1A, 1B**).

**Figure 1.**
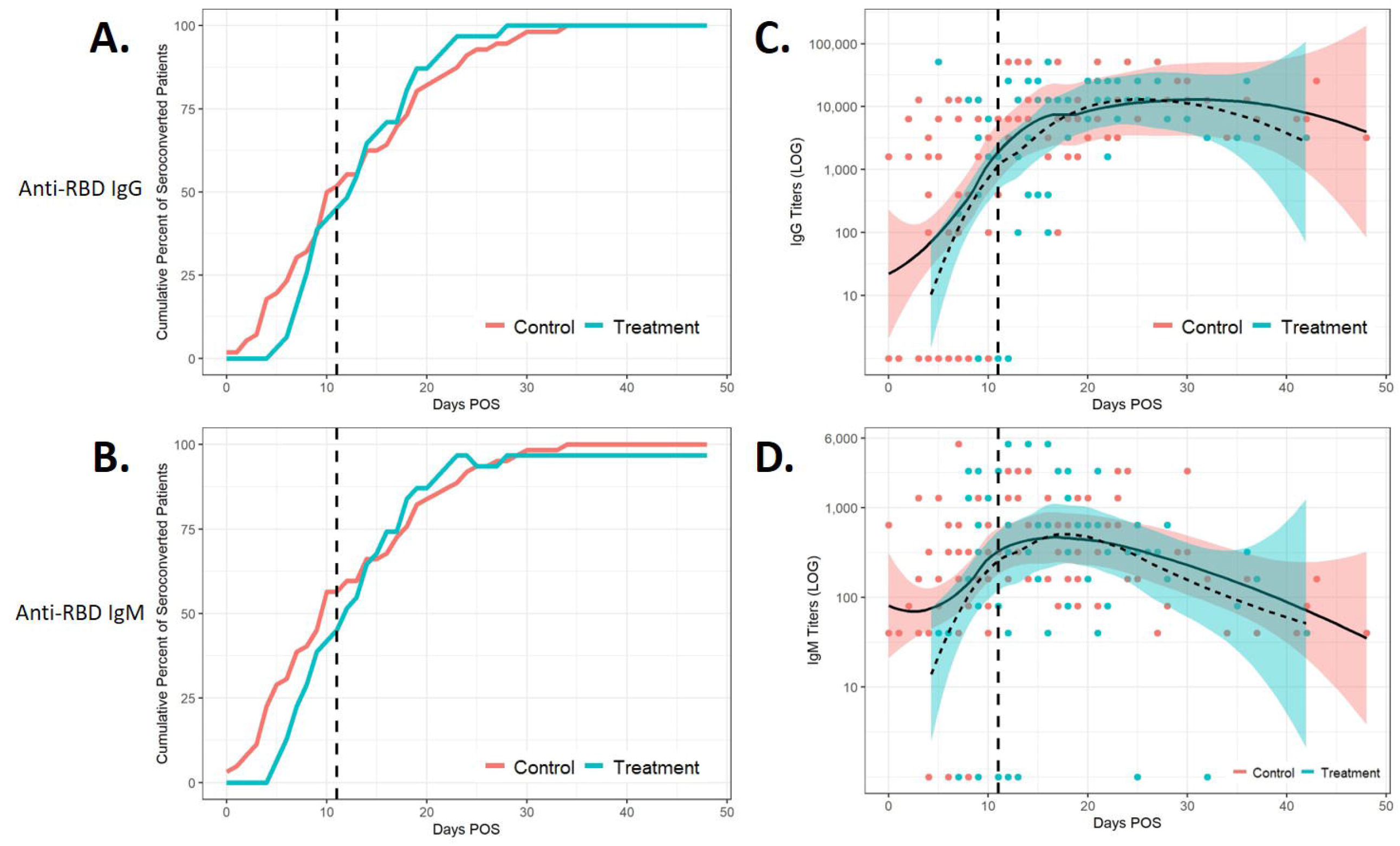
Kinetics of anti-RBD IgG and anti-RBD IgM response in COVID-19 patients as a function of number of days post-onset of symptoms. Cumulative frequency, as a percent of seropositive CCP, transfused patients (celeste blue line) and non-CCP patients (pink line), were plotted against the number of days post-onset of symptoms (POS). Seropositive was defined as any titer measurement of IgG (**1A**) or IgM (**1B**) greater than 1:100 and 1:40, respectively. Patients were assumed to be seronegative prior to the first measurement (**1A-B**). Scatter plots to model IgG (**1C**) and IgM (**1D**) antibody titer trends over days POS are indicated by Loess regression curves; a span of 0.75 and shadings indicate the 95% confidence intervals (CI) for CCP recipients (dotted line, blue CI) overlaid with the curves for control patients (solid line, pink CI). Vertical dashed line represents median days POS at which transfusion occurred. All titer levels were converted to a log 10 scale.

□The longitudinal profiles of anti-RBD IgG and IgM responses were analyzed using a LOESS regression model. The anti-RBD IgG response peaked between 20-30 days POS and slowly decayed thereafter for both CCP recipients and controls (**Figure 1C**). The anti-RBD IgM response peaked between 15-25 days POS and rapidly decayed thereafter for both CCP recipients and controls (**Figure 1D**).

Lastly, the kinetics of individual patients’ anti-RBD IgG and IgM response were compared between CCP recipients (**Figure 2A-B**) and controls (**Figure 2C-D**). As was observed at the overall population level in **Figure 1**, the kinetics of the anti-RBD IgG and IgM responses were similar in both CCP recipients and control groups at the individual patient level (**Figure 2A-D**). The respective responses were higher for patients on ECMO (red) compared to those solely mechanically ventilated (green), and solely mechanically ventilated subjects had higher titers than non-ventilated patients (blue) (**Figure 2A-D**).

**Figure 2.**
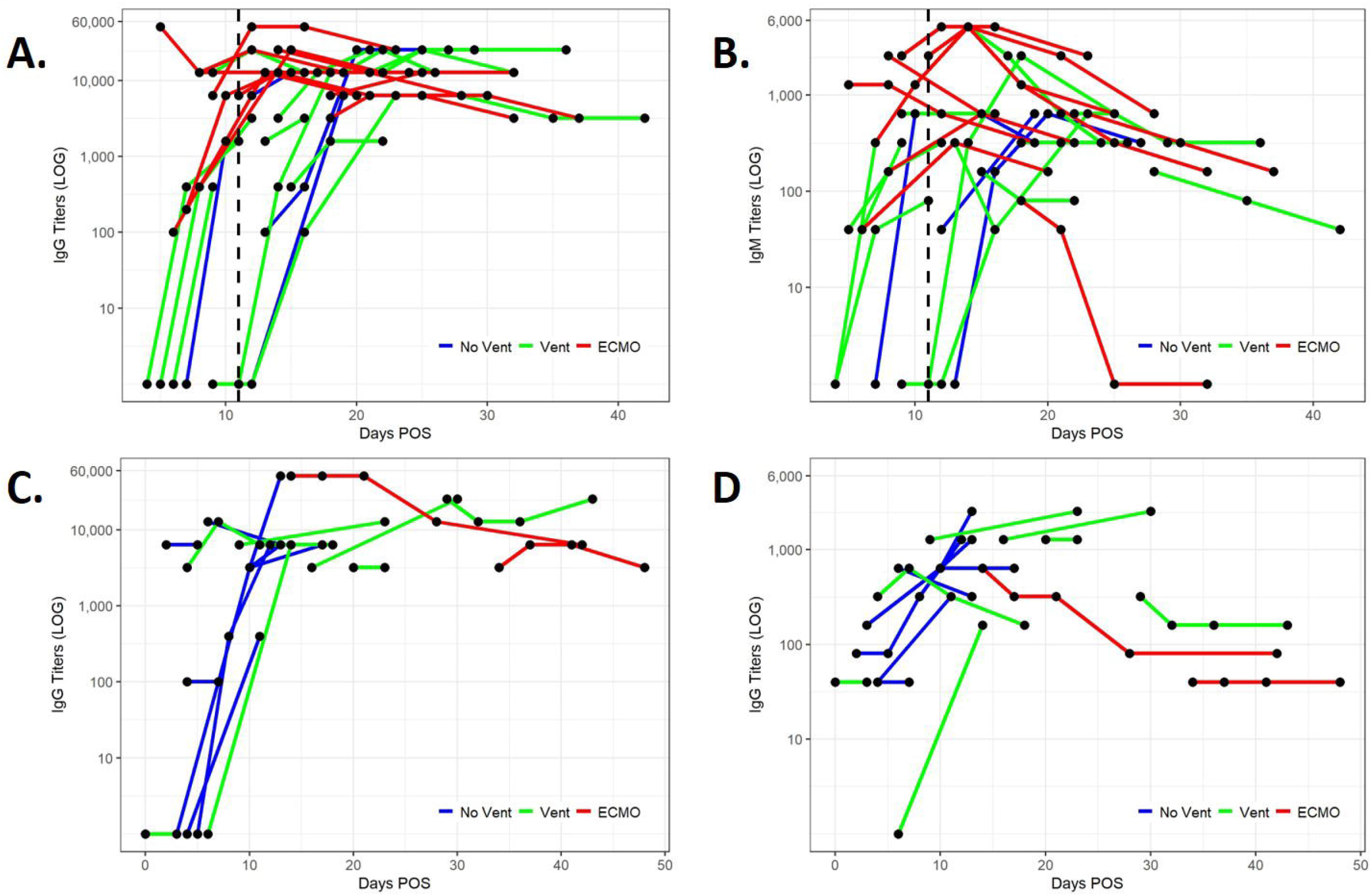
Anti-RBD IgG and anti-RBD IgM longitudinal responses of individual COVID-19 patients over time post-onset of symptoms. Anti-RBD IgG (**2A**) and IgM (**2B**) longitudinal responses for individual CCP recipients (n=31/31). The first data point of each line represents patient antibody titer immediately prior to CCP transfusion, and subsequent dots representing titers on post-transfusion days (3,7 and 14). Only controls with sequential data points are shown (n=18/68) (**2C-D**). Individual controls anti-RBD IgG (**2C**) and IgM (**2D**) longitudinal titer responses are shown on each line with each dot for sequential days post-onset of symptoms. CCP and control patient samples are stratified based on the level of respiratory support needed; no ventilation (dark blue), ventilation only (green), ECMO (red) (**2A-D**). The vertical dashed line represents median days POS at which transfusion occurred. All titer levels were converted to a log 10 scale.

Immediately prior to CCP transfusion, patients median anti-RBD IgG and IgM titers were 1:3200 (IQR, 1:50 to 1:9600) and 1:320 (IQR,1:40 to1: 640), respectively. Additionally, preceding CCP administration, eight out of 31 (25.8%) patients were anti-RBD IgG seronegative (Median 6.5, IQR 4.75 to 9.5 days POS) (**Figure 2A**), six out of 31 (19.3%) had no detectable anti-RBD IgM (Median 8, IQR 4.75 to 10.5 days POS) (**Figure 2B**), and five out of 31(16%) were seronegative for both (Median 7, IQR 4 to 9 days POS). Interestingly, three out of these five CCP recipients seronegative for both anti-RBD IgG and IgM died within 30 days of transfusion, one of whom was a recent kidney transplant recipient on immunosuppressive therapy. In the control group, 16 out of 68 (23.5%) were anti-RBD IgG seronegative at the time of initial sample collection (Median 5, IQR 3.75 to 6.25 days POS), five out of 68 (7.4%) had no detectable anti-RBD IgM (Median 6, IQR 6 to 6 days POS), and four out of 68 (5.9%) were negative for both (Median 6, IQR 6 to 6.5 days POS).

### Anti-RBD IgG and IgM response distribution in CCP recipients is similar in control patients

Next, we compared the distribution of anti-RBD IgG (**Figure 3A**) and IgM (**Figure 3B**) responses between recipients and controls depending on the level of respiratory support needed. Anti-RBD IgG and IgM titers in non-ventilated CCP recipients were similar to non-ventilated controls (IgG Median titers: 1:6400 and 1:3200 (**Figure 3A**), (IgM Median titers: 1:480 and 1:160 (**Figure 3B**)).

**Figure 3.**
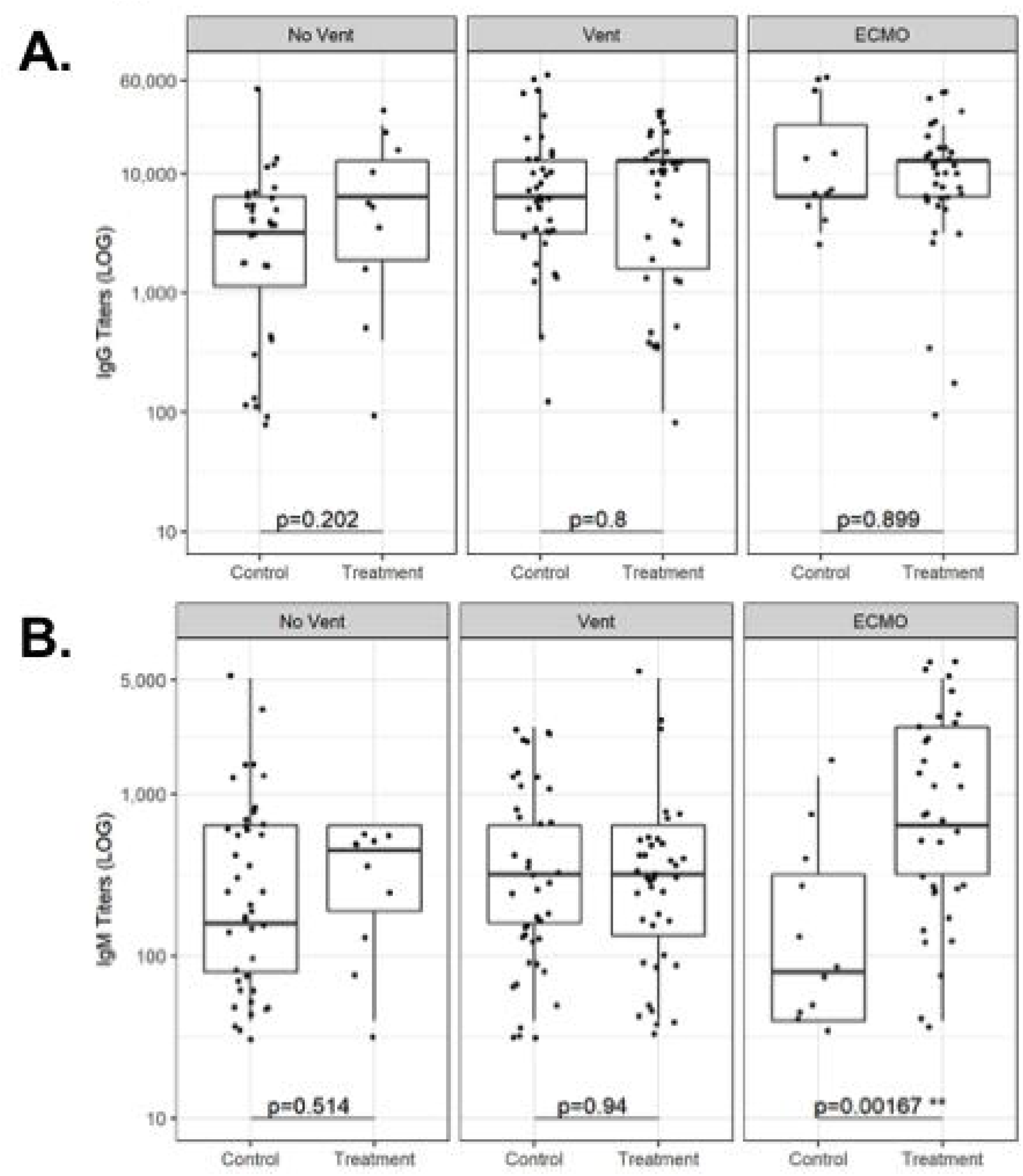
Anti-RBD IgG and anti-RBD IgM response distribution in COVID-19 patients stratified by disease severity and respiratory support needed. Distribution of anti-RBD IgG (**3A**) and IgM (**3B)** titers in CCP patients compared to controls depending on the three levels of respiratory support needed. Seronegative samples were excluded (**3A-B**). Statistical analysis was performed using a Kruskal-Wallis test. An alpha value of 0.05 or less was considered statistically significant. All titer levels were converted to a log 10 scale.

Mechanically ventilated CCP recipients had similar anti-RBD IgG and IgM titers compared to ventilated control patients (IgG median titers: 1:12800 and 1:6400 (**Figure 3A**), (IgM median titers: 1:320 and 1:320 (**Figure 3B**).

CCP recipients on ECMO had similar anti-RBD IgG titers compared to control patients on ECMO (median titers: 1:12800 and 1:6400 (**Figure 3A**). In contrast, IgM titer levels were higher in CCP recipients versus control patients (1:640 and 1:80, respectively (**Figure 3B**). The observed increase in anti-RBD IgM titers in CCP recipients on ECMO may be due to a difference in the number of days POS at which samples were drawn. IgM measurements were taken at a median of 18 days POS for CCP recipients compared to 28 days POS for control patients (p=0.015) (data not shown).

### Clinical outcomes of CCP recipients are similar to matched control COVID-19 patients

CCP recipients and matched control COVID-19 patients (Control B) presented with similar thirty-day in-hospital mortality (**Figure 4A**). When stratifying the two groups based on disease severity, no difference in thirty-day in-hospital mortality was observed (**Figure 4B-C**). Additionally, CCP recipients and matched controls were similar in their respective median ICU LOS and median hospital LOS (**Table 2**). The subgroups of CCP recipients also had similar ICU LOS and hospital LOS when compared to their respective matched control subgroups (**Table 2**). CCP recipients and matched controls also had a similar median number of days on mechanical ventilation and median duration on ECMO (**Table 2**).

**Table 2:**
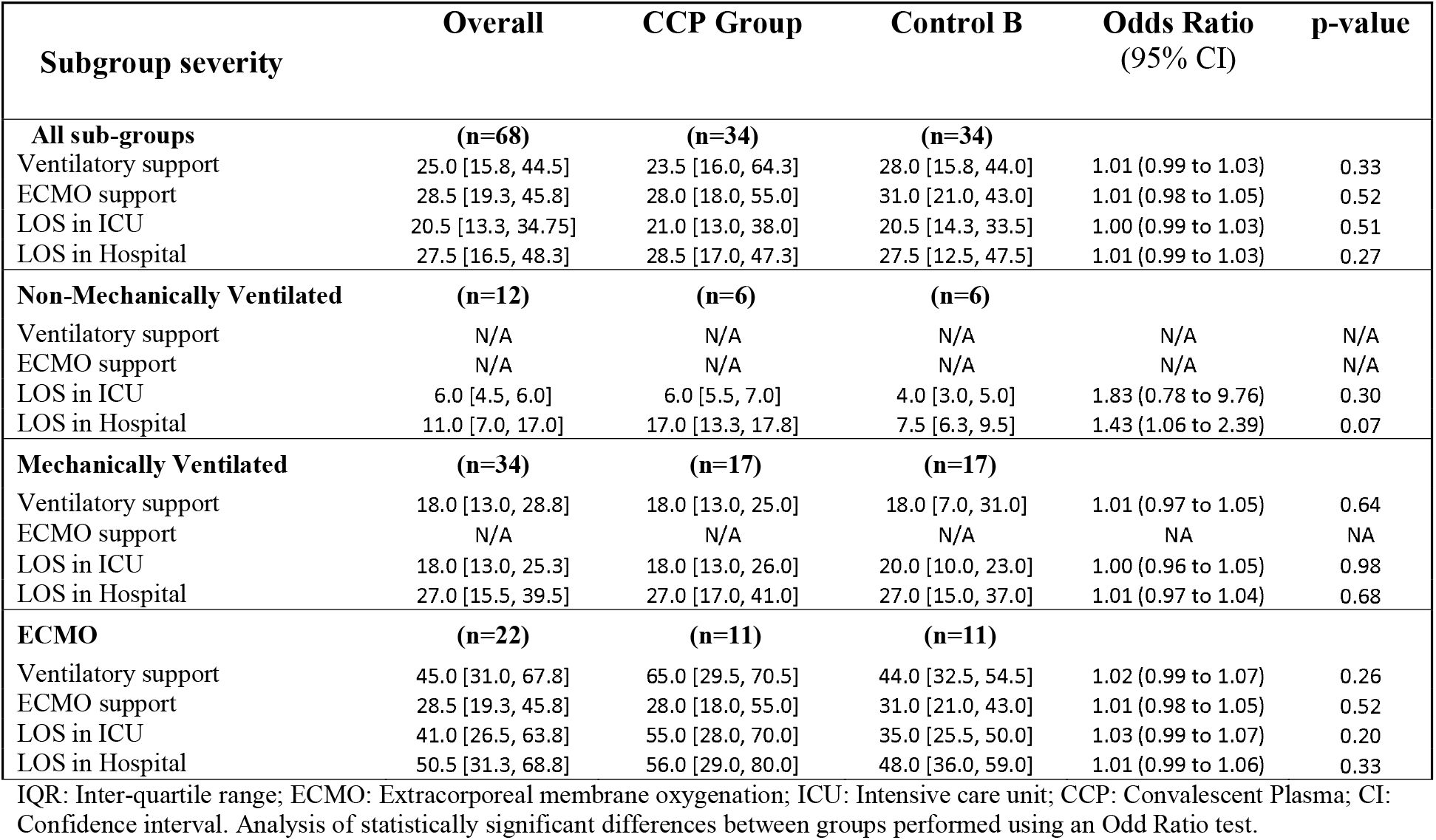
Comparison of secondary clinical outcomes in subgroups of COVID-19 severity. Values are number of days as median [IQR].

**Figure 4.**
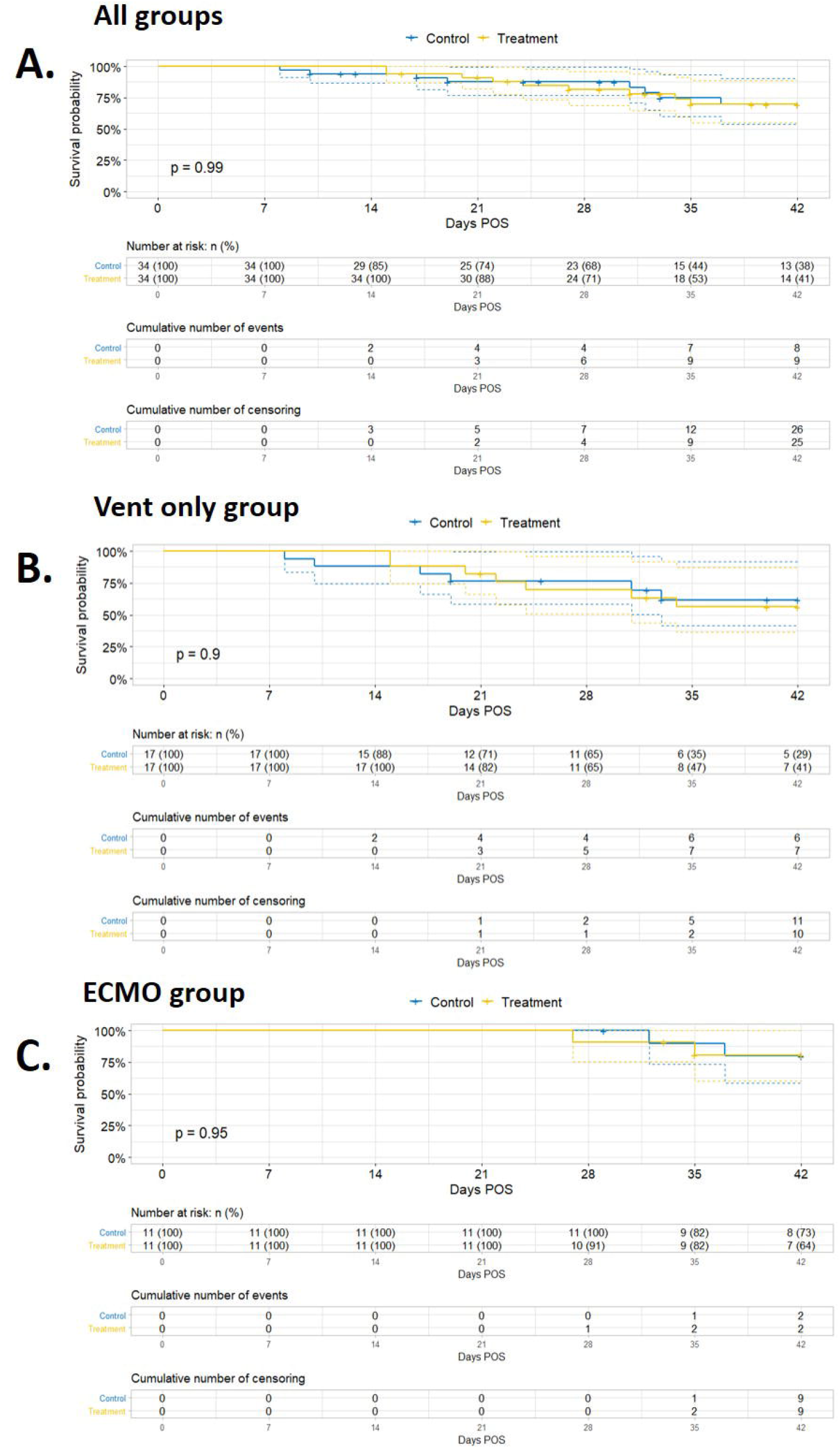
Thirty-day mortality of hospitalized COVID-19 patients. CCP transfused patients (yellow line) compared to matched controls (blue line) with Kaplan-Meier curves. Patients and controls were considered censored at discharge or at 42 days POS. All CCP recipients compared to all control patients (**4A**). CCP recipients on ventilator alone compared to control patients on ventilator alone (**4B**). CCP recipients on ECMO compared to control patients on ECMO (**4C**). Statistical analysis performed using a log-rank test. An alpha value of 0.05 or less was considered statistically significant.

Lastly, CCP recipients had a greater 3-day decline in plasma CRP levels following administration of CCP compared to controls matched for day of symptoms (**Table 3**). There were no other differences in changes in inflammatory marker concentrations when comparing treatment and matched control groups in this study.

**Table 3:** Comparison of changes in inflammatory marker levels.

| **Change in**<br>**Concentration**<br>**median (IQR)** | **Overall**<br>**(n=68)** | **CCP Group**<br>**(n=34)** | **Control B**<br>**(n=34)** | **Odds Ratio**<br>**(95% CI)** | **p-value** |
| --- | --- | --- | --- | --- | --- |
| **CRP mg/dL** |  |  |  |  |  |
| Day 3-Day 0 | 2.3 [-13.4, 2.9] | -3.7 [-20.2, 0.6] | 0.40 [-3.00, 13.40] | 0.95 (0.90 to 0.99) | 0.023 |
| Day 7-Day 0 | -6.0 [-17.7, 1.3] | -8.5 [-19.3, 0.2] | -2.10 [-14.90, 6.80] | 0.98 (0.93 to 1.02) | 0.32 |
| Day 14-Day 0 | -10.3 [-16.8, -1.8] | -10.3 [-25.0, 4.4] | -10.85 [-16.42, -4.62] | 1.01 (0.97 to 1.06) | 0.557 |
| **D-dimer (ng/mL)** |  |  |  |  |  |
| Day 3-Day 0 | -60.0 [-2105.0, 420.0] | -295.0 [-1910.0, 392.5] | -40.0 [-4370.0, 600.0] | 1.00 (1.00 to 1.00) | 0.964 |
| Day 7-Day 0 | -230.0 [-2440.0, 1948.0] | -465.0 [-2437.5, 1736.0] | 440.0 [-3185.0, 3630.0] | 1.00 (1.00 to 1.00) | 0.984 |
| Day 14-Day 0 | 545.0 [-2097.5, 2648.3] | 750.0 [-140.00, 6,647.5] | -1780.0 [-7363.0, 1870.0] | 1.00 (1.00 to 1.00) | 0.076 |
| **Ferritin (ng/mL)** |  |  |  |  |  |
| Day 3-Day 0 | -46.1 [-249.5, 75.0] | -46.8 [-278.1, 82.0] | -45.3 [-174.2, 56.6] | 1.00 (1.00 to 1.00) | 0.923 |
| Day 7-Day 0 | -39.9 [-491.3, 224.5] | -200.3 [-568.5, 129.9] | 96.4 [-11.8, 293.5] | 1.00 (1.00 to 1.00) | 0.97 |
| Day 14-Day 0 | -192.6 [-520.2, 15.3] | -435.8 [-586.3, -7.3] | -147.2 [-359.8, 395.7] | 1.00 (1.00 to 1.00) | 0.5 |
| **Fibrinogen (mg/dL)** |  |  |  |  |  |
| Day 3-Day 0 | -15.0 [-95.0, 153.0] | -44.0 [-155.5, 126.0] | 81.0 [-15.0, 153.0] | 1.00 (1.00 to 1.00) | 0.097 |
| Day 7-Day 0 | 23.0 [-190.0, 107.0] | -26.5 [-264.0, 86.8] | 52.0 [23.0, 179.0] | 1.00 (0.99 to 1.00) | 0.121 |
| Day 14-Day 0 | -147.0 [-347.0, 8.5] | -229.0 [-383.0, -38.0] | -111.5 [-188.3, 49.3] | 1.00 (0.99 to 1.00) | 0.357 |

SD: Standard deviations; CRP: C-reactive protein; CCP: Convalescent Plasma; CI: Confidence Interval. Analysis of statistically significant differences between groups performed using an Odds Ratio Test.

## DISCUSSION

The kinetics of SARS-CoV-2 IgG and IgM antibodies from plasma of COVID-19 patients transfused with CCP were comparable to those from a cohort of COVID-19 patients who did not receive CCP. Furthermore, most CCP recipients already had detectable SARS-CoV-2 IgG antibodies in their plasma prior to transfusion, which occurred at a median of 11 days following symptom onset. The highest SARS-CoV-2 IgG antibody titers were observed in the plasma of the sickest subgroup of patients requiring both ventilatory and ECMO support. CCP recipients compared to a matched control group did not show any mortality benefit at 30 days post-transfusion, nor a reduction in either ICU or hospital LOS, or duration of mechanical ventilation / ECMO support; besides, with stratification based on disease severity, no effect on mortality between the two groups was observed. Interestingly, there was a decline in CRP inflammatory marker at day 3 following CCP transfusion compared to control levels.

### Comparison with other studies

While some of the current findings corroborate results from earlier studies, others contradict them. Hergorova et al. reported a modest survival benefit in a matched control study in patients with severe COVID-19 following CCP transfusion within seven days of hospitalization(10). Salazar et al. from Houston found that in a prospective, propensity score-matched study, patients transfused with CCP within 72 hours of admission experienced the most benefit compared to the control group(11). By contrast, in an open-label, randomized controlled trial conducted in India, CCP was not associated with a reduction in overall mortality or progression to severe COVID-19(12), even when administered within three days of symptom onset. A retrospective study in China at the beginning of the pandemic with ten patients showed improved oxygenation and better patient survival following CCP transfusion(13). However, four out of the 10 patients had high (≥1:640) SARS-CoV-2 neutralizing antibody titers prior to CCP transfusion, suggesting that the patient’s own immunity may have been responsible for the recovery rather than CCP transfusion. Although these data show mixed results, they support prioritization of CCP transfusion to COVID-19 patients who present early within 3-5 days of symptom onset when native antibody production is still in the fledgling stages, or in those patients who are immunosuppressed (e.g., hypogammaglobulinemia)(14). In the current study, three of the CCP recipients who were seronegative prior to transfusion died within 30 days, one of these was a kidney transplant recipient who was receiving T-cell immunosuppression prior to COVID-19 diagnosis, suggesting that T cell response may also be important for controlling SARS-CoV-2 during the acute phase of the infection.

Although a difference in the rise of antibody titers could have been expected in CCP recipients around day 15 post symptom onset (approximately day three post-transfusion), the increase in IgG and IgM titers was similar in both treated and control groups. These data are consistent with reports showing the general COVID-19 patient population, detectable IgG, and IgM in plasma between four and seven days post-onset of symptoms(15). In a randomized control trial, PlasmAr Study, of 215 patients with severe pneumonia, total SARS-CoV-2 antibody titers were higher in the CCP treated group at day two post-transfusion. Still, no effect on 30-day clinical outcome and mortality between treated versus placebo groups was observed (16). Similarly, in the current study, most patients treated with CCP presented with severe COVID-19, and over 80% of them were intubated at the time of transfusion. The strength of the antibody response was greater in patients on mechanical ventilation and/or on ECMO than in non-ventilated patients; this observation is consistent with a report by Klein et al. and others, showing an association between COVID-19 disease severity and antibody titer levels(17–19).

### Strengths and limitations of this study

This study adds to the current literature on CCP efficacy by characterizing the kinetics of SARS-CoV-2 antibodies following CCP transfusion, which has not been previously described longitudinally in comparison to control plasma samples from non-transfused COVID-19 patients. Furthermore, the study reports on CCP therapeutic responses in specific subgroups of patients who require either solely ventilatory support and/or ECMO support. Our institution is a referral center for those most severe cases of COVID-19 in the state of Maryland. Thus, we believe that the data presented can provide generalizability on the effect of CCP transfusion in subgroups of critically ill patients presenting with different levels of respiratory support.

To strengthen the study, we compared the antibody titer measurements by ELISA to those obtained on a commercially available instrument, the Ortho VITROS total anti-SARS-CoV-2 Ig platform, which had been previously validated against a SARS-CoV-2 neutralizing live-cell assay(20, 21). The median IgG titers prior to CCP transfusion were high (>1:3200). Interestingly, Salazar et al. showed that anti-RBD IgG titers greater than 1:1350 correlated with SARS-CoV-2 neutralization (VN) titers greater than 1:160 at 80% probability(20). VN titer > 1:160 is the recommended level by the FDA in CCP products using the Ortho VITROS IgG platform at a signal to a cut-off ratio (S/C) of 12. While we were not able to compare our titer results directly to the Ortho VITROS IgG platform, Luchsinger et al. showed that both the Ortho VITROS total Ig and IgG platforms, set at a median S/C values of 101 and 11.7 respectively, correlated well to neutralizing antibody results and gold-standard ELISAs(21). Our validation showed that the median anti-RBD IgG titer of 1:6400 in 24 control samples, also tested by the Ortho VITROS total Ig method, showed a median S/C of 490 for total anti-SARS-CoV-2 Ig, suggesting that titers of ≥1:6400 and 1:3200 on the ELISA used in the present study are much higher than the recommended S/C of 12 and are indicative of high neutralizing antibody titers.

There are limitations associated with this study. Although the blood supplier qualified the CCP donations as high tittered (>1:160), the exact titers were not provided, thus we cannot certify that the CCP units transfused in the study have a titer greater than 1:160. The study is also limited by the fact that it is an observational study; thus, its reliability in examining clinical outcomes compared to a prospective, randomized, control trial is not as robust; but at the advent of the first surge of the pandemic, a randomized trial was not practical at our institution. Lastly, the numbers of patients enrolled in each group are small, but the clinical outcomes of CCP recipients were compared to matched control patients hospitalized at the same hospital. This approach decreased bias due to the clustering of enrolment.

### Policy implications

The use of convalescent plasma as a treatment for Covid-19 is authorized in the United States under an Emergency Use Authorization with no specific guidance on prioritization of particular patient subgroups based on disease severity. CCP resources are limited, and the current data further guides decision-making about CCP transfusion to critically ill patients with COVID-19.

## Conclusion

In conclusion, the current data may further guide clinicians in defining eligibility criteria for CCP transfusion for the treatment of COVID-19. Indeed, these data do not support CCP transfusion to patients with severe COVID-19, especially if presenting with plasma SARS-CoV-2 IgG and IgM neutralizing antibody levels at presentation. Taken together with the current literature, our findings confirm that CCP is probably most effective when administered to patients with low antibody titers, who are earlier in the disease course, and who do not yet have complicated COVID-19.

## Data Availability

Upon request Data sources can be provided for results presented in the study

## ACKNOWLEDGMENTS

We thank Dr. Wilbur Chen for facilitating the procurement of the plasmids for the RBD production. We also thank all the staff of the Transfusion Services at each University of Maryland Medical Systems hospital. We thank the staff and physicians at both the American Red Cross and New York Blood Center for their coordinated efforts in providing timely convalescent plasma units to our patients.

We thank for their continuous support Dr. Christenson, Director of the Core Chemistry and Immunology Laboratory; Dr. Barreuta, Chief Medical Officer at the University of Maryland Upper Chesapeake Medical Center; Joanne Marshall, Supervisor, Office of Corporate Research Compliance University of Maryland Medical System.

## CONTRIBUTORSHIP STATEMENT

MNK and EW completed the experimental work and clinical data collection and wrote the manuscript. HB, CD, AMH, AT, and JB completed the clinical data collection and reviewed and edited the manuscript, PZ completed all statistical analysis and editing of the manuscript. EMB contributed to the design and editing of the manuscript. KEM and MJF designed, supervised the experimental work, and wrote the manuscript. All authors have reviewed, edited, and approved the manuscript before submission. MJF is the guarantor.

## TRANSPARENCY STATEMENT

As the senior author and corresponding author, I, Magali J Fontaine, affirm that the manuscript is an honest, accurate, and transparent account of the study being reported; that no significant aspect of the study has been omitted; and that any discrepancies from the study as initially planned have been explained.

## ETHICAL APPROVAL

The study was approved by the University of Maryland Baltimore Institutional Research Subject review board (Protocol HP-00092606).

## DATA SHARING

Upon request, Data sources can be provided for the results presented in our study.

## FIGURE LEGENDS

**Supplemental Figure 1.**
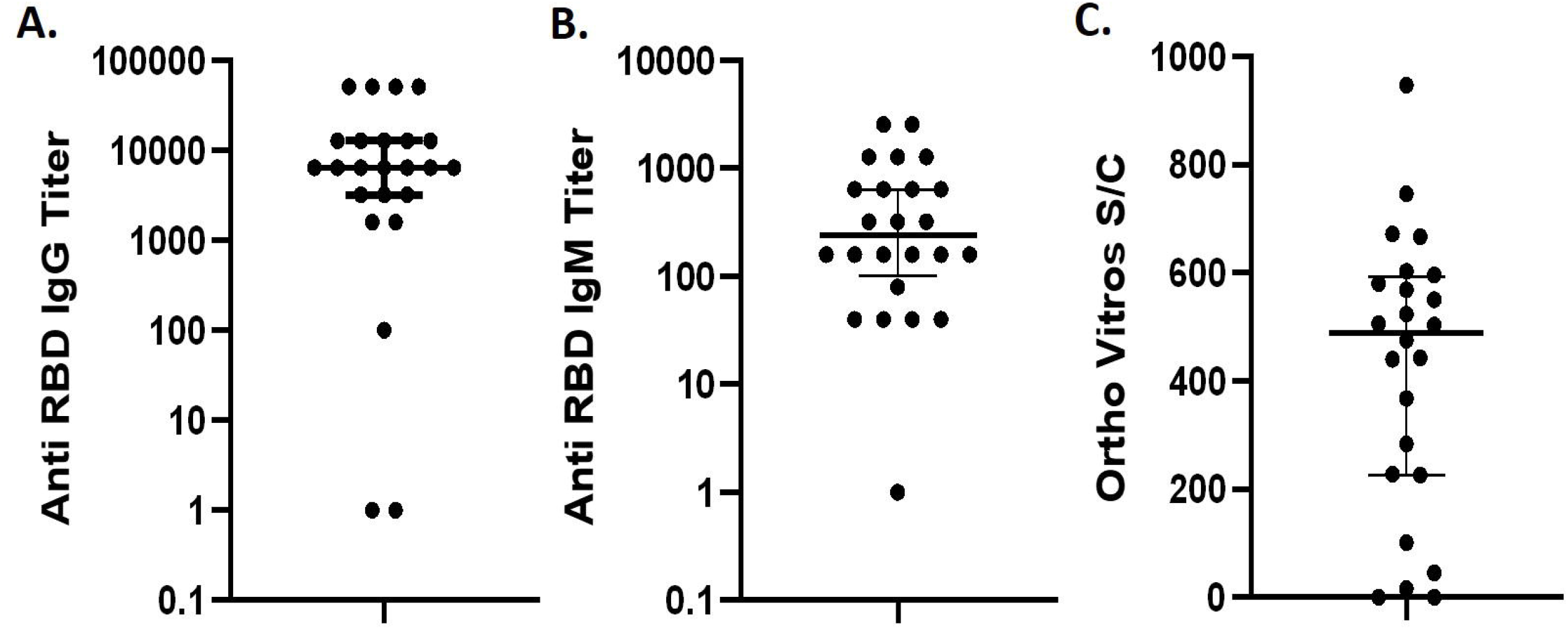
Antibody response comparisons using different platforms. Anti-RBD IgG (**S1A**) and IgM (**S1B**) titers as well as the signal cut-off (S/C) from the Ortho VITROS total anti-SARS-CoV-2 Ig platform (**S1C**) of 24 COVID-19 control patients. IgG titers below 1:100, IgM titers below 1:40, and S/C values below 1.0 were considered negative. Bars represent the median and interquartile range (IQR). All titer levels were converted to a log 10 scale (**S1A-B**).

